# A Whole-Brain Functional Connectivity Model of Alzheimer’s Disease Pathology

**DOI:** 10.1101/2021.01.13.21249597

**Authors:** Ruchika S. Prakash, Michael R. McKenna, Oyetunde Gbadeyan, Anita R. Shankar, Rebecca Andridge, Douglas W. Scharre

**Affiliations:** Department of Psychology, The Ohio State University, OH, USA; Division of Biostatistics, The Ohio State University, OH, USA; Department of Neurology, Division of Cognitive Neurology, The Ohio State University Wexner Medical Center, OH, USA

**Keywords:** Whole-brain functional connectivity, machine learning, Alzheimer’s disease, global cognition

## Abstract

Early detection of Alzheimer’s disease (AD) is a necessity as prognosis is poor upon symptom onset. Although previous work diagnosing AD from protein-based biomarkers has been encouraging, cerebrospinal (CSF) biomarker measurement of AD proteins requires invasive lumbar puncture, whereas assessment of direct accumulation requires radioactive substance exposure in positron emission tomography (PET) imaging. Functional magnetic resonance imaging (fMRI)-based neuromarkers, offers an alternative, especially those built by capitalizing on variance distributed across the entire human connectome. In this study, we employed connectome-based predictive modeling (CPM) to build a model of functional connections that would predict CSF p-tau/Aβ_42_ (PATH-fc model) in individuals diagnosed with Mild Cognitive Impairment (MCI) and AD dementia. fMRI, CSF-based biomarker data, and longitudinal data from neuropsychological testing from the Alzheimer’s Disease NeuroImaging Initiative (ADNI) were utilized to build the PATH-fc model. Our results provide support for successful in-sample fit of the PATH-fc model in predicting AD pathology in MCI and AD dementia individuals. The PATH-fc model, distributed across all ten canonical networks, additionally predicted cognitive decline on composite measures of global cognition and executive functioning. Our highly distributed pathology-based model of functional connectivity disruptions had a striking overlap with the spatial affinities of amyloid and tau pathology, and included the default mode network as the hub of such network-based disruptions in AD. Future work validating this model in other external datasets, and to midlife adults and older adults with no known diagnosis, will critically extend this neuromarker development work using fMRI.

**Significance Statement:** Alzheimer’s disease (AD) is clinical-pathological syndrome with multi-domain amnestic symptoms considered the hallmark feature of the disease. However, accumulating evidence from autopsy studies evince support for the onset of pathophysiological processes well before the onset of symptoms. Although CSF- and PET-based biomarkers provide indirect and direct estimates of AD pathology, both methodologies are invasive. In here, we implemented a supervised machine learning algorithm – connectome-based predictive modeling – on fMRI data and found support for a whole-brain model of functional connectivity to predict AD pathology and decline in cognitive functioning over a two-year period. Our study provides support for AD pathology dependent functional connectivity disturbances in large-scale functional networks to influence the trajectory of key cognitive domains in MCI and AD patients.

## Introduction

According to recent estimates by the Alzheimer’s Association, 5.8 million Americans are currently facing AD, with 13.8 million older adults projected to be diagnosed by 2050 (1). This has resulted in significant investment from the National Institutes of Health (NIH) towards understanding the etiology, progression, and treatment of Alzheimer’s disease (2, 3). Cognitive deficits are considered hallmark symptoms of AD, with notable impairment to episodic memory; however, other sub-domains of cognitive functioning are also degraded including executive dysfunction, processing speed, and attentional control (4–7). There is unequivocal evidence from imaging and autopsy studies that the neurodegenerative process in AD, especially alterations in fluid-based and imaging-based biomarkers, initiates well before, and in some instances, without the presence of known cognitive symptoms (4–7). Additionally, autopsy studies also confirm the absence of AD neuropathology in 10-30% of individuals diagnosed to have AD-related dementia by experts (8), questioning the specificity of the prototypical amnestic clinical syndrome for AD neuropathology. Thus, a research framework recently proposed by the National Institute on Aging has emphasized a paradigm shift in conceptualizing AD away from clinical symptoms to a biologically driven approach, defined by either postmortem or in vivo examination of fluid or imaging biomarkers (2). Furthermore, with no current treatment to reverse the effects of the disease, the prognosis of AD is poor after the appearance of symptoms, making early detection a critical necessity.

Previous research has offered encouraging results in using both CSF-based and imaging-based biomarkers to categorize between healthy controls and individuals with varying stages of AD (9–12). Two well-studied biomarkers in AD are CSF levels of Aβ_42_, one of the most reliable diagnostic isoforms of Aβ, and CSF levels of hyperphosphorylated tau (p-tau), which is thought to be more specific to AD pathology than other measures such as total-tau (13). Of these CSF-based markers, there is increasing support for a ratiometric measure of p-tau/Aβ_42_, combining variance across the two critically implicated proteinopathies of amyloidosis and tauopathy, to have one of the best diagnostic accuracies (14–16). For example, Hansson et al.(14) comparing the accuracy rates for the recently developed and reliable Elecsys-based immunoassays of Aβ42, t-tau/Aβ42, and p-tau/Aβ42, provided support for the two ratiometric measures to have stronger concordance with amyloid PET than CSF-Aβ_42_ alone. With new radiotracers available for imaging of tau depositions, there is also recent growth in the use of PET to identify imaging-based diagnostic biomarkers of AD pathology (17). However, the invasive nature of repeated lumbar punctures, and the introduction of radioactive substances during PET, necessitate the use of other noninvasive imaging techniques, such as fMRI to develop neuromarkers that reliably predict symptom onset, disease progression, and demonstrate sensitivity to change as a function of key therapeutic agents. Like biomarker accumulation, changes in functional connectivity occur well before the onset of clinical symptoms (3, 18), thus positioning functional connectivity as an effective way to predict future cognitive decline.

fMRI is an imaging modality that indirectly captures neuronal activity by modeling the hemodynamic response. Much of the prior research examining functional connectivity changes within AD focused on understanding activity- and connectivity-based disruptions in the default mode network (19), a set of midline cortical regions with early metabolic and histopathological changes as a result of AD (20, 21). However, increasingly, there has also been interest in examining the intra-network changes in other key canonical networks. Although studies repeatedly demonstrate disruptions in intra-network connectivity of large-scale networks supporting cognitive processes, like the frontoparietal network, salience network, and the dorsal attention network, there are significant differences across studies regarding the networks that show disruptions (22, 23). Similarly, there are also discrepancies reported in the literature on the extent and pattern of inter-network disconnections seen as a function of aggregation of amyloid and tau proteins, with some studies suggesting reduced anticorrelations between networks with increasing pathology (24–26), and other studies attributing the inter-network dysfunction to aging rather than AD pathogenesis (23). These discrepancies, across studies, thus highlight the limitations of group-based approaches that neither account for the continuous and heterogeneous nature of AD neurodegeneration, nor differentiate between network level changes occurring due to aging compared to those occurring due to neurodegenerative pathologic processes. Furthermore, with recent incorporation of machine learning in analyses of neuroimaging data, the field has witnessed a significant shift in our reliance on investigating one or two canonical networks of the brain, to conceptualizing complex psychiatric and neurological disorders, including AD, as byproducts of systemic changes in the entire connectome (27–29). Connectome-based predictive modeling (CPM) is one such whole-brain, data-driven approach that allows for the identification of connectivity patterns within the individual’s entire connectome that are predictive of disease states, symptom severity, or cognitive functioning (30, 31). In what is now considered a set of formative studies in the development of functional MRI-based models, Rosenberg and colleagues, derived and tested the generalizability of a whole-brain, FC-based model of sustained attention: saCPM (30, 32–34). The saCPM using leave-one-out cross-validation procedure predicted over 70% of variance in sustained attention in a cohort of young adults on whom the model was derived (30). More importantly, the saCPM neuromarker was then generalized to predict attention-based deficiencies in children with attention-deficit/hyperactivity disorder (30); predict cognitive performance on other metrics of attention, like inhibitory control (32) and executive control (33); and serve as a surrogate biomarker for treatment efficacy with increased connectivity following methylphenidate treatment for ADHD (30). Extending this saCPM to older adults, our lab, in collaboration with Rosenberg and colleagues, recently evinced support for the saCPM neuromarker to predict attentional control in healthy older adults, with the saCPM neuromarker explaining 38.4% of the variance in Stroop task performance across a sample of young adults and older adults (34).

Following the success of the saCPM, there has been a growing interest in the use of CPM to predict symptom severity in neurodegenerative populations (35) as well as in psychiatric populations (36, 30, 37, 38). Within AD, Lin et al.(35) is the only study, to our knowledge, examining the utility of CPM in predicting baseline levels of cognitive functioning in the ADNI sample. They found that the increased connectivity within the frontoparietal network and increased connectivity between the frontoparietal network and default mode network were related to worse baseline cognitive scores on the ADAS11 measure of overall cognitive functioning.

However, no study, to our knowledge, has yet utilized this approach to explore links between whole-brain patterns of functional connectivity and underlying biological measures in AD, specifically CSF-based biomarkers. Here, we employed the CPM method to develop a whole-brain marker of functional connectivity representing CSF-based AD pathology. Specifically, we used the ratiometric measure of p-tau/Aβ_42_ to develop a connectome-based model of FC in symptomatic older adults and further determined the in-sample fit of this model in predicting decline over a period of two years in global cognition, memory functioning, and executive functioning in the same sample.

## Results

### Distribution of AD Pathology and Cognitive Composites

We included 62 participants with MCI and 21 participants with AD in the final sample. As shown in Table 1, participants across the two groups were predominantly White (91.65%), with an average age of 72 years, and education of 16 years. 14.3% of MCI individuals and 50.8% of AD individuals were APOE ε4 positive. Scores on all three of the cognitive composites declined over time (see Table 2), with the largest declines seen for PACC, followed by ADNI-MEM, and ADNI-EF.

**Table 1.**
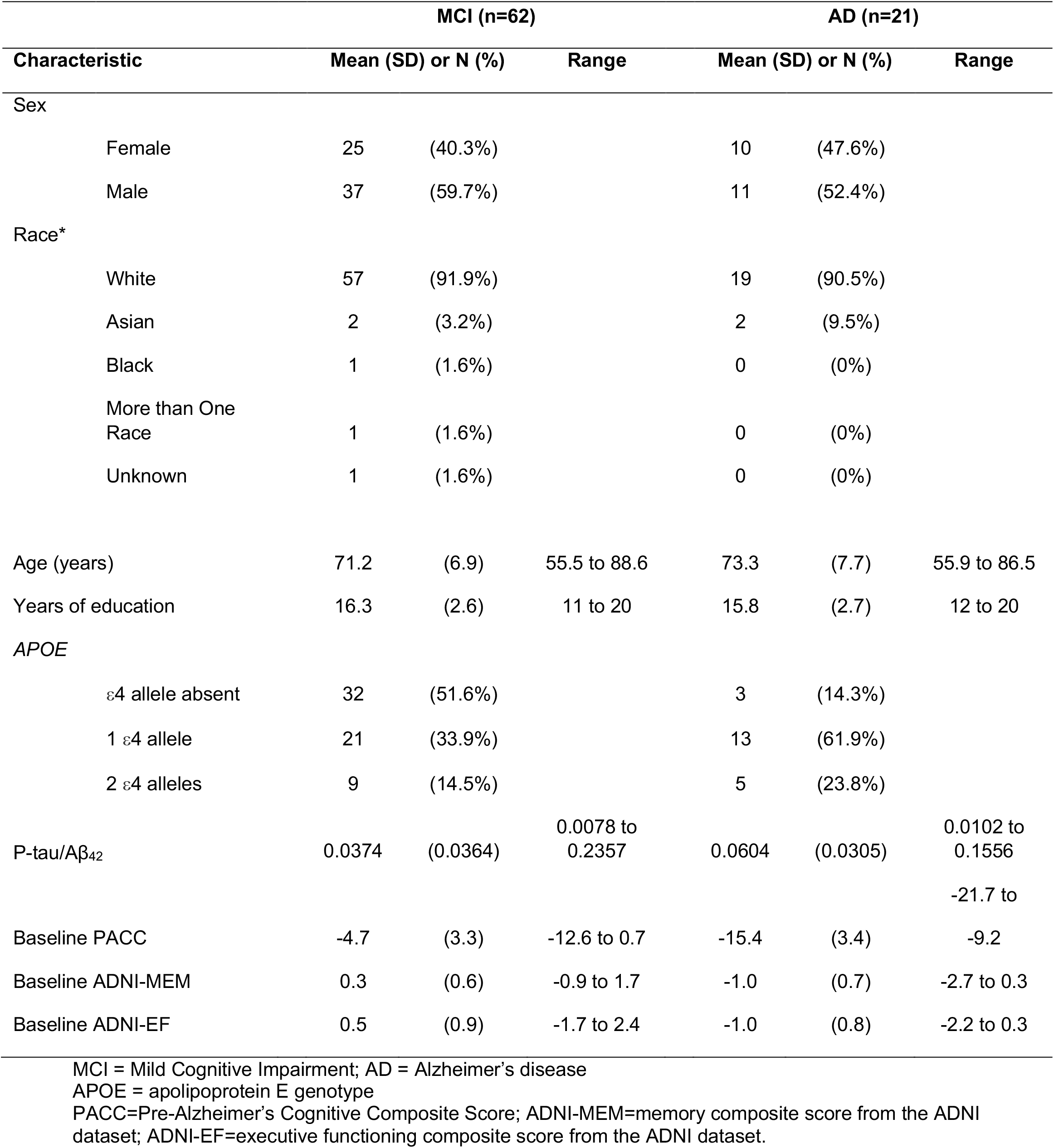
Baseline characteristics of participants by diagnostic groups.

| Characteristic | MCI (n=62) |  |  | AD (n=21) |  |  |
| --- | --- | --- | --- | --- | --- | --- |
|  | Mean (SD) or N (%) |  | Range | Mean (SD) or N (%) |  | Range |
| Sex |  |  |  |  |  |  |
| Female | 25 | (40.3%) |  | 10 | (47.6%) |  |
| Male | 37 | (59.7%) |  | 11 | (52.4%) |  |
| Race* |  |  |  |  |  |  |
| White | 57 | (91.9%) |  | 19 | (90.5%) |  |
| Asian | 2 | (3.2%) |  | 2 | (9.5%) |  |
| Black | 1 | (1.6%) |  | 0 | (0%) |  |
| More than One Race | 1 | (1.6%) |  | 0 | (0%) |  |
| Unknown | 1 | (1.6%) |  | 0 | (0%) |  |
| Age (years) | 71.2 | (6.9) | 55.5 to 88.6 | 73.3 | (7.7) | 55.9 to 86.5 |
| Years of education | 16.3 | (2.6) | 11 to 20 | 15.8 | (2.7) | 12 to 20 |
| APOE |  |  |  |  |  |  |
| ε4 allele absent | 32 | (51.6%) |  | 3 | (14.3%) |  |
| 1 ε4 allele | 21 | (33.9%) |  | 13 | (61.9%) |  |
| 2 ε4 alleles | 9 | (14.5%) |  | 5 | (23.8%) |  |
| P-tau/Aβ<sub>42</sub> | 0.0374 | (0.0364) | 0.0078 to 0.2357 | 0.0604 | (0.0305) | 0.0102 to 0.1556 |
| Baseline PACC | -4.7 | (3.3) | -12.6 to 0.7 | -15.4 | (3.4) | -21.7 to -9.2 |
| Baseline ADNI-MEM | 0.3 | (0.6) | -0.9 to 1.7 | -1.0 | (0.7) | -2.7 to 0.3 |
| Baseline ADNI-EF | 0.5 | (0.9) | -1.7 to 2.4 | -1.0 | (0.8) | -2.2 to 0.3 |

MCI = Mild Cognitive Impairment; AD = Alzheimer's disease
APOE = apolipoprotein E genotype
PACC=Pre-Alzheimer's Cognitive Composite Score; ADNI-MEM=memory composite score from the ADNI dataset; ADNI-EF=executive functioning composite score from the ADNI dataset.

**Table 2.** Estimated slopes (change in outcome for 1 month increase in time) over two years for cognitive outcomes from linear mixed effects models

| Measure | Overall (n=83) |  |  | MCI (n=62) |  |  | AD (n=21) |  |  |
| --- | --- | --- | --- | --- | --- | --- | --- | --- | --- |
|  | Slope | (SE) | p-value | Slope | (SE) | p-value | Slope | (SE) | p-value |
| PACC | -0.12 | (0.021) | **<.0001** | -0.07 | (0.022) | **0.004** | -0.29 | (0.050) | **<.0001** |
| ADNI-MEM | -0.0086 | (0.0024) | **0.0006** | -0.007 | (0.0025) | **0.009** | -0.014 | (0.0059) | **0.02** |
| ADNI-EF | -0.015 | (0.0036) | **<.0001** | -0.006 | (0.0039) | 0.11 | -0.042 | (0.0087) | **<.0001** |

### Resting-State Connectivity Patterns Predict AD Pathology

We employed connectome-based predictive modeling (see Figure 1) to develop a whole-brain functional connectivity marker of AD pathology. Employing the leave-one-out-cross-validation technique, connectome-based models were trained to predict pathology score for the test participant during each round of the cross-validation. The respective predicted pathology scores were correlated with observed pathology scores to index model performance. This resulted in two distinct sub-networks: 1) a high pathology network, representing a set of edges correlated positively with levels of CSF-biomarker accumulation, and 2) a low pathology network, representing a set of edges negatively associated with pathology. As the CSF-based ratio of p-tau/Aβ_42_ quantifies the concentrations of the two proteinopathies in the CSF, with elevated levels representing higher pathological load, we only examined the high pathology network – PATH-fc model in this study. Moreover, as we note below, the low pathology network, did not reach statistical significance and resulted in a total of 2 edges, thus corroborating our *a priori* decision of not including the low pathology network in our final model. We found that the overall CPM model successfully predicted AD pathology scores (*r*_*s*_ = 0.32, *p* = 0.028, 1000 iteration permutation testing). Additionally, the high pathology model, the PATH-fc model, comprising of 581 edges (Figure 2A & B), successfully predicted AD pathology (*r*_*s*_ = 0.25, *p* = 0.013, 1000 iteration permutation testing; Figure 2C). However, the predictability of AD pathology in the low pathology model did not reach statistical significance (*r*_*s*_ = 0.07, *p* = 0.27, 1000 iteration permutation testing). The low pathology model comprised of 2 edges. Thus, in all subsequently reported results, we focus only on the high pathology model.

**Figure 1.**
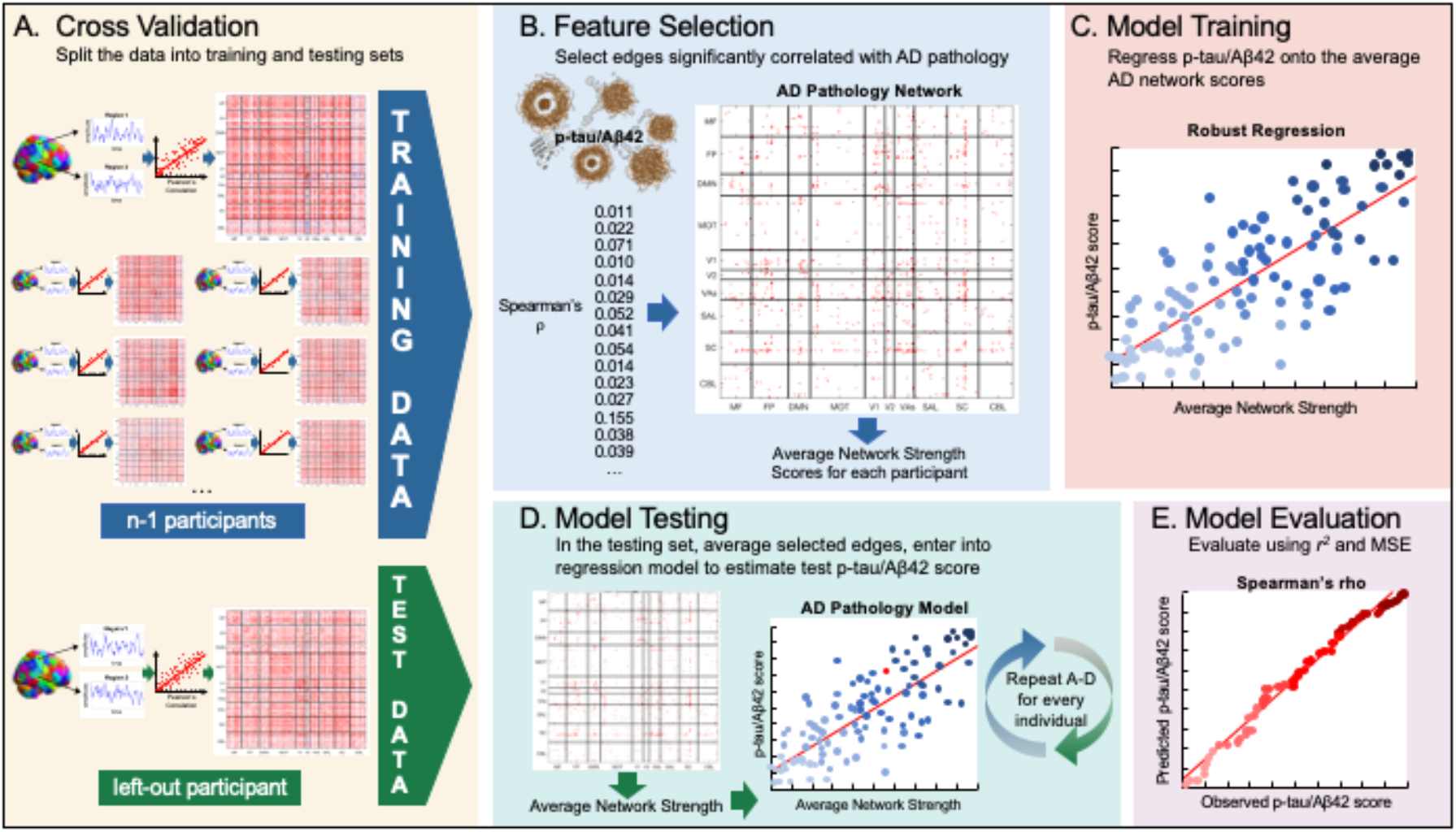
Schematic Presentation of CPM Model.

**Figure 2.**
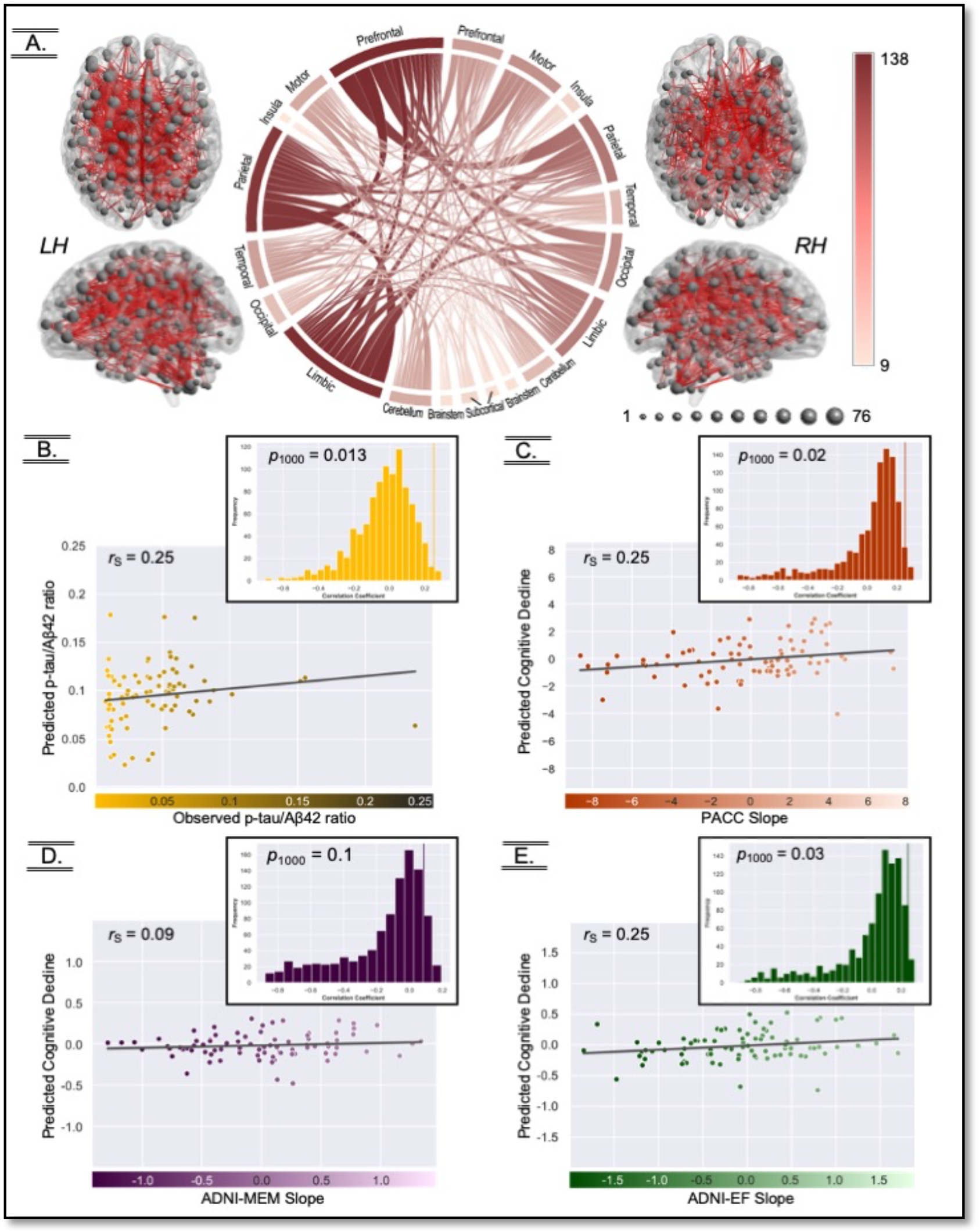
Visual depiction of the PATH-fc model and the in-sample fit for AD pathology and cognitive decline. A. Presents the anatomical distribution of the PATH-fc model. The 581 edges included nodes in all the macroscale regions, with the left hemisphere showing greater involvement than the right. B. Internal model fit of the PATH-fc model to the ADNI pathology data. C-E. Internal model fit of the PATH-fc model to predict cognitive decline in PACC, ADNI-MEM, and ADNI-EF, respectively. The scatterplots represent Spearman correlation between observed scores (AD pathology, PACC, ADNI-MEM, and ADNI-EF) and predicted scores for the same metric derived using cross-validation. Histogram represents the randomly shuffled connectivity matrices and respective metric pairings over 1000 iterations to compute the final p-value for the significance.

### PATH-fc Model is Predictive of Cognitive Decline

Having established that resting-state functional connectivity predicts AD pathology, we next examined whether the PATH-fc model would predict decline in cognitive functioning in MCI and AD individuals. To address this aim, the CPM pipeline was modified to predict the rate of cognitive decline using high pathology model strength. To evaluate the performance of the PATH-fc model in predicting cognitive decline, we then correlated the predicted and observed rates of cognitive decline over a two-year period on PACC, ADNI-MEM, and ADNI-EF. We found that models trained on PATH-fc strength were able to significantly predict PACC slope (*r*_*s*_ = 0.25, *p* = 0.02) and ADNI-EF slope (*r*_*s*_ = 0.25, *P* = 0.03), but not ADNI-MEM slope (*r*_*s*_ = 0.09, *p* = 0.1). Figure 2C presents the correlation plots demonstrating the fit between predicted and observed cognitive decline. Similar results were observed when PATH-fc strength scores were correlated with observed rates of cognitive decline (Supplementary Figure S3A).

### PATH-fc is a Highly Distributed Model, and Primarily Features Inter-Network Edges

PATH-fc is a highly distributed model, with the 581 edges of the model distributed across all ten macroscale regions (see Materials and Methods). 483 edges were in the left hemisphere and included 179 ipsilateral connections and 304 contralateral connections. In contrast, 402 edges included were in the right hemisphere, with 98 ipsilateral connections. Within the left hemisphere, nodes with the greatest number of edges were localized to the prefrontal cortex, the parietal cortex, and the limbic. In the right hemisphere, the nodes of the PATH-fc model were more distributed within the parietal cortex, the limbic regions, and the occipital cortex.

Additionally, the 581 edges of the PATH-fc model also included inter- and intra-network edges from nearly all of the 10 canonical networks (Figure 3A). The motor network, default mode network, and frontoparietal network were the three most commonly represented networks within the PATH-fc model (Figure 3A). However, to account for network size in the Shen parcellation scheme, we normalized edge counts using the following formula, as described previously (39):

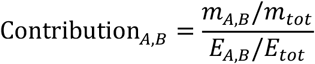

where Contribution_*A,B*_ represents the relative contribution of edges between networks A and B, *m*_*A,B*_ is the number of edges between A and B in our model, *m*_*tot*_ is the total number of edges in our model, *E*_*A,B*_ is the number of possible edges between A and B, and *E*_*tot*_ is the total number of possible edges in the whole brain. As shown in the matrix in Figure 3B, accounting for network size demonstrates the relatively high contribution of the default mode network and reduces the prominence of the motor network. The top three networks by relative contribution are the default mode (mean contribution=1.97), visual association (left middle occipital gyrus; mean contribution=1.40), and frontoparietal (left superior frontal gyrus; mean contribution=1.30) networks.

**Figure 3.**
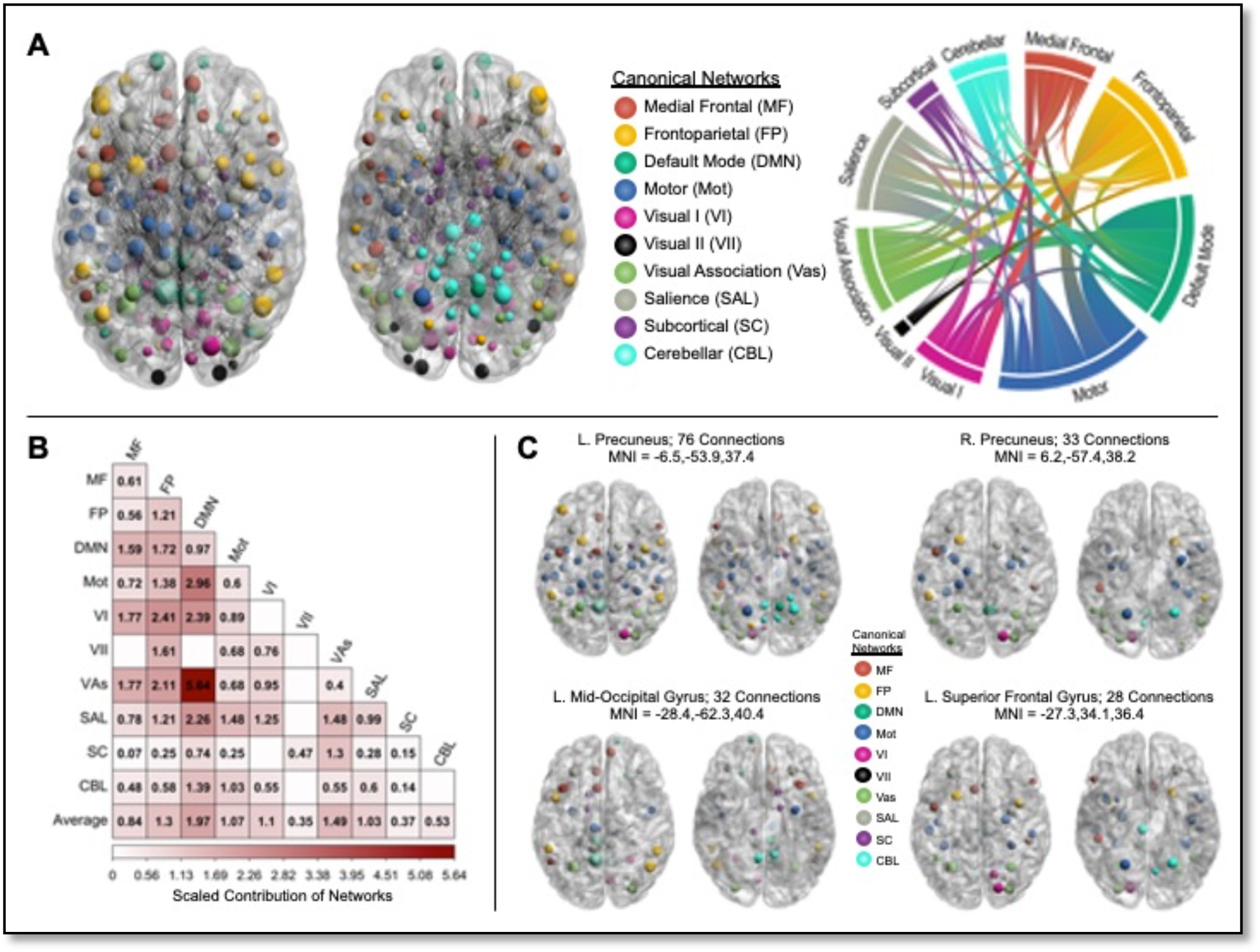
Network localization of the PATH-fc model. A. Presents the involvement of the 10 canonical networks in the PATH-fc model, with nodes parcellated based on Noble et al. (2019).Ribbons in the ring plot visualization are proportional to the representation of each network in the PATH-fc model. The matrix presented in panel B shows the relative contribution of each network to the PATH-fc model adjusted using the formula described in Greene et al., 2019. Panel C highlights the anatomical and network localization of the four high degree nodes in the PATH-fc model, with two nodes belonging to the default mode network, one to the salience network, and one the visual association network.

Additionally, we were interested in examining if PATH-fc primarily featured intra-network or inter-network connections. Of the 581 edges, 39 were intra-network edges and 542 edges were inter-network connections. Applying the same contribution formula to these edges shows a higher relative contribution of inter-network edges compared to intra-network edges (average inter-network contribution = 1.051, average intra-network contribution: 0.5959).

Outlier analysis indicated that four nodes had an elevated number of edges within PATH-fc (Figure 3C). The top two nodes with the most edges in PATH-fc were nodes in the left and right posterior cingulate cortex (PCC) (left MNI coordinates = −6.5, −53.9, 37.4; right MNI coordinates = 6.2, −57.4, 38.2); major hubs in the default mode network. 18.76% of edges in PATH-fc included one of these two nodes. The third and fourth nodes with the most edges were in the visual association (MNI coordinates: −28.4, −62.3, 40.4) and salience (MNI coordinates − 27.3, 34.1,36.4) networks.

### AD Pathological Models are Robust to Key Confounds

When using additional edge-selection threshold values of 0.1, 0.05,0.005, and 0.001 (40), we observed that model performance was comparable across all thresholds (Supplementary Table S1). Replicating our analyses with an additional functional brain atlas (Glasser360), we found that high pathology model successfully predicted AD pathology (Supplementary Figure S2A). We next examined whether this newly derived high pathology model would also predict rates of cognitive decline in a similar fashion as our main analysis. We found a significant prediction for PACC scores (Supplementary Figure S2B) and a marginally significant prediction for ADNI-MEM scores (Supplementary Figure S2C). However, the prediction for ADNI-EF scores did not reach statistical significance (Supplementary Figure S2D). We also observed that the correlations of PATH-fc predicted AD pathology scores or PATH-fc network strength with the estimated rates of decline on PACC and ADNI-EF, but not ADNI-MEM, were significant (Supplementary Figure S3B). Taken together, these findings suggest that the PATH-fc model was robust to potential confounds.

## Discussion

The goal of this study was to leverage methodological developments in the field of computational neuroscience to derive a whole-brain marker of synchronous neural activity using CSF-based biomarkers of AD pathology. Using connectome-based predictive modeling that captures variance distributed across the brain (32, 33), we derived the PATH-fc model – a highly distributed network of edges with representation across the various macroscale regions – to predict variance in the ratio of p-tau/Aβ_42_ in MCI and AD individuals. The PATH-fc model, consistent with existent literature on the spatial distribution of amyloid (21, 41) and tau pathology (42) in MCI and AD individuals, included all ten canonical networks, with nodes in the default mode network, the frontoparietal network, and the visual association network showing the highest degree of representation. Critically, the PATH-fc model, in addition to predicting AD pathology, predicted the rates of cognitive decline, notably on measures of global cognition and executive functioning. This study is the first to employ computational models of functional connectivity to build a pathology-based model with implications for cognitive decline in AD.

Aggregation of extra-cellular amyloid plaques and intra-cellular neurofibrillary tangles containing hyperphosphorylated tau are the defining pathological features of AD (2, 43, 44). CSF- and PET-based biomarkers of Aβ and tau have been reliably found to differentiate between healthy and pathological aging (45–47), predict conversion of MCI individuals to AD (48), and explain variance in clinical symptoms of dementia (49). However, the accumulation of Aβ-plaques and neurofibrillary tangles follow differentiated spatiotemporal trajectories, with amyloid biomarkers representing one of the earliest signs of AD neuropathological changes (50, 51), and accumulation of tau inclusions a necessary pathological marker for AD diagnosis (2). Critically, of the various biomarkers of AD-based pathology, the CSF-based ratio of p-tau/Aβ_42_ provides one of the best diagnostic accuracies among fluid- and PET imaging-based biomarkers (52, 53), shows the largest effect sizes, compared with Aβ_42_ or p-tau alone in distinguishing between cognitively normal, MCI, and AD individuals (54), and predicts performance on the Clinical Dementia Rating Scale (55), measures of driving performance (56), and neuropsychological-based measures of global cognition, memory, executive functioning, and semantic fluency (54, 57). As such, in our study, we employed this combined ratio of p-tau/Aβ_42_ to derive a whole-brain, functional connectivity marker of AD pathology that would allow us to capture the distinct spatial affinities of the two proteinopathies. Although it would be interesting to also examine networks predicting Aβ_42_ and p-tau separately, we were interested in deriving one connectome-based marker that would comprehensively capture the disruptions in large-scale functional networks because of the synergistic effect of accumulating β-amyloid and tau. Our results, demonstrating successful in-sample fit of the PATH-fc model in predicting p-tau/Aβ_42_, provide support for the use of computational modeling techniques in deriving functional connectivity-based models for biological signatures of AD pathology.

The PATH-fc model, derived using a synergistic metric of the two hallmark AD proteinopathies, is a distributed model that includes representation from all macroscale regions and the ten canonical networks. As this model was derived in MCI and AD individuals, these results are consistent with *in vivo* and *in vitro* studies that implicate the involvement of large swaths of the neocortex in AD pathogenesis, especially in the later stages of the disease when clinical symptoms have already manifested (58–62). The deposition of AD pathology, specifically tau depositions, is hypothesized to spread in a prion-like manner, with pathology often originating in densely connected nodes of the human brain, like the PCC/precuneus and the entorhinal cortices, and spreading trans-neuronally to other highly functionally connected regions, thus engulfing many of the large canonical networks (63–65). Confirming the involvement of large-scale functional networks, the PATH-fc model included 581 edges, with the frontoparietal, default mode, motor, visual I, visual association, and salience networks showing large contributions relative to their network size in the overall model.

Interestingly, a qualitative examination of the involvement of various canonical networks in the PATH-fc model, provides a striking overlap with the spatial affinities of amyloid and tau pathology, which include the densely connected nodes of the default mode network (41, 63) and the entorhinal cortices (64, 66) respectively. Specifically, PET studies have shown the aggregation of misfolded amyloid proteins to begin in the nodes of the default mode network, with early stages including the mid-cortical posterior and anterior structures involving the PCC, the precuneus, the ACC, and the medial orbitofrontal gyri, and advanced stages including the limbic and the subcortical regions. Tau deposits, on the other hand, appear to originate in the transentorhinal cortices, with pathology spreading in a prion-like manner to areas of the perirhinal cortex, including the hippocampus, the limbic areas, and then widely to the association cortices. These spatial affinities of Aβ and tau-based tangles to target distinct brain regions also mirrors the findings of multi-modal investigations that associate regional binding of PET-based tracers to disruptions in resting-state functional connectivity of large-scale canonical networks (63, 67–70). For example, Elman et al. (63) examined the associations between global and regional PiB retention in cognitively normal older adults and functional connectivity of the canonical networks during rest. Both global and regional PiB retention was associated with reduced within-network connectivity of nodes in the default mode network, with amyloid pathology also influencing the connectivity of other large-scale networks supporting exogenous attention, like the dorsal attention network and the frontoparietal network. Moreover, they also showed reduced anti-correlations between nodes of the frontoparietal and dorsal attention network and the precuneus, suggesting that the disruptions caused by AD pathology extend beyond intra-network connections, and influence the connectivity of the default mode network with other networks of the brain.

Our results, consistent with the existent literature, highlight the nodes of the default mode network in the PATH-fc model, with both intra-network and inter-network connections of the default mode network predicting p-tau/Aβ_42_ ratio. This result also aligns with other studies examining the association between CSF-based measures of AD pathology and functional connectivity, such that lower levels of Aβ_42_ and p-tau are associated with decreased connectivity within nodes of the default mode network, but also extends to interactions between the default mode network and the salience, frontoparietal, and language networks (71). Our study, however, goes beyond establishing correlations between CSF-based measures of AD pathology and functional connectivity, and generates a predictive marker of whole-brain functional connectivity that critically implicates the default mode network as the hub of AD pathology-based disruptions in functional connections. Moreover, two of the four nodes showing the highest degree – the number of connections a node has with other nodes – were in the default mode network and included both the left and right precuneus. Pathological and PET-based studies further show that these regions are hubs of metabolic activity (72, 73), demonstrate aerobic glycolysis (74, 75), and show regional reductions in glucose metabolism (76, 77), thus potentially making them particularly vulnerable to deposition of AD pathology.

Interestingly, the PATH-fc model, in addition to capturing the involvement of the default mode network, also implicated the involvement of several other networks, most notably the frontoparietal and the visual association networks, with connections between the default mode network and the visual association cortices showing the highest relative contribution to the PATH-fc model. These results support previous PET-fMRI and animal studies implicating the association cortices as potentially key regions involved in the trans-neuronal spread of AD pathology likely via their shared neuronal activity. *In-vitro* animal studies evince support for the spreading of pathogenic tau via axonal connections to anatomically connected, yet spatially remote regions rather than adjacent regions (58, 59). Similarly, human PET-fMRI studies show that the pattern of functional connectivity is associated with tau covariance such that regions with high levels of tau show high functional coupling and those with low regions of tau depositions are coupled with other nodes with low accumulation (67). Importantly, this covariance between functional connectivity and tau deposition was independent of the Eucledian distance between the regions, suggesting that tau propagation was dependent on the synaptic connectivity rather than spatial adjacency. This covariance between tau spread in highly connected regions is also independent of overall functional connectivity of the cortex given that tau accumulation is inversely associated with overall functional connectivity (68), thus suggesting that higher neuronal coupling of spatially distant, yet connected regions results in spread of AD pathology from cell-to-cell in large-scale structurally and functionally connected networks.

Collectively, the results of our study and other human multi-modal investigations, highlight the covariance between accumulation of pathological proteins and disruptions of synchronous neuronal activity within and between canonical networks. In fact, it is widely appreciated that these disruptions antedate the appearance of clinical symptoms, and although there are equivocal findings with regards to associations between individual pathological markers and clinical symptomatology, combined metrics of amyloidosis and tauopathy are more consistently associated with declining cognitive functioning. Within the ADNI dataset, we have demonstrated that p-tau/Aβ_42_ ratio predicts the two-year slope of cognitive decline on composite measures of global cognition, executive functioning, and episodic memory in 619 individuals with MCI and 229 older adults with AD (54). Extending the current literature evincing support for associations between pathological and clinical metrics of AD, in this study, we demonstrate that the PATH-fc model derived to predict p-tau/Aβ_42_ ratio can also successfully predict the change in cognitive functioning, specifically on measures of global cognition and executive functioning. These results, although cross-sectional in nature, imply that our computational model of pathology-dependent functional connectivity predicts changes in measures of global cognition and executive functioning in MCI and AD individuals over the course of two years. Thus, although previous studies have demonstrated associations between disruptions in functional connectivity and cognitive functioning, and pathology-based biomarkers of AD pathology and cognition, our study provides support for AD pathology dependent functional connectivity disturbances in large-scale functional networks to influence the trajectory of key cognitive domains in MCI and AD patients.

It is important to consider the results and interpretations of this study in the context of certain limitations. First, to derive a model of AD pathology, we employed the CSF-based p-tau/Aβ42 ratio, with this metric representing a summary measure of fluid-based pathology. This ratiometric measure, however, lacks the spatial specificity that could be derived from PET-based markers of AD pathology, thus resulting in a connectome model that may be qualitatively different from one generated using regionally-specific PET amyloid and tau ligands. Although not ideal, CSF-based markers are more readily available in publicly available longitudinal datasets; have strong diagnostic accuracies (14–16); and have been associated with aberrant FC in large-scale canonical networks (78). Additionally, CPM methodology is able to identify connections that are most important for linking FC to the desired behavior or characteristic, thus identifying connections between nodes that are linked to this global summary measure of pathology. Future studies, integrating across PET and fMRI, may be able to create joint connectome models that concatenate regional update of PiB and tau tracers with functional connectivity estimates to build spatially-specific pathology-based models.

Second, although the PATH-fc model predicted the rate of cognitive decline over a two-year period, because of attrition in the ADNI dataset that differed as a function of diagnostic group, we did not include more longitudinal follow-up assessments. To validate the model for predicting clinically meaningful cognitive decline, it would be important to demonstrate the utility of this connectome-based model in predicting conversion of diagnostic status, either from cognitively normal to MCI, or from MCI to AD. As such, although there is promise in this pathology-dependent functional connectivity marker for cognitive functioning of MCI and AD individuals, it would be critical to extend this model beyond the current results.

And, finally, our study was predominantly cross-sectional in nature, and although we used computational predictive modelling to derive a pathology-dependent marker, longitudinal investigations would be needed to assess the impact of pathology progression on large-scale networks.

Overall, in the last 10 years, great strides have been made in our understanding of this clinical-pathological neurodegenerative disease, with clear consensus that sensitive and specific biomarkers are critically needed for refining diagnostic accuracy, predicting progression of the disease, and identifying preventative and therapeutic treatments. Our results provide support for use of functional connectivity measures to build a model that is predictive of hallmark AD pathology and cognitive consequences of the structural and functional alterations. Future studies, validating this model in external datasets to predict pathology and conversion of diagnostic status, are needed to determine the utility of this model.

## Materials and Methods

### Participants and Experimental Design

The current study is a secondary analysis of publicly available data from The Alzheimer’s Disease Neuroimaging Initiative (ADNI). ADNI is a multicenter study designed to assess AD progression, collecting longitudinal, multi-modal data involving MRI, PET, CSF- and plasma-based biological markers, and detailed neuropsychological measures. Inclusionary criteria for participants in the ADNI dataset involved the following requirements: aged between 55-90 years, overall good health condition, fluent in either English or Spanish, and geriatric depression scale score of less than six. We used the ADNIGO and ADNI2 phases since ADNI1 and ADNI3 do not have matching MRI parameters, and ADNI3 is still being collected. 200 individuals with MCI were collected as part of ADNIGO, with an additional 400 AD pathology individuals (150 AD, 250 MCI) collected as part of ADNI2. However, resting-state data was only collected at a limited number of sites, leaving 100 (49% Female) participants who had baseline fMRI data: 76 are individuals with mild cognitive impairment (MCI), and 24 are individuals with AD.

Diagnostic criteria for classifying participants on the spectrum of pathological aging at baseline involved a combination of subjective reports, neuropsychological assessment, and physician assessment (79). Specifically, for MCI participants, the following criteria were employed: 1) MMSE score ≥ 24, 2) scoring within education-adjusted range for the Logical Memory II subscale (EMCI: 9-11 for 16+ years of education, 5-9 for 8-15 years of education, 3-6 for 0-7 years of education; LMCI: ≤8 for 16+ of education, ≤4 for 8-15 years of education, ≤2 for 0-7 years of education), 3) a Clinical Dementia Rating score of 0.5, 4) self or partner reported memory complaint(s), and 5) absence of AD dementia or any other neurological condition. AD participants were required to meet the following criteria: 1) MMSE score between 20-26, 2) scoring below education-adjusted cut-offs for the Logical Memory Scale II (8 for 16+ of education, ≤4 for 8-15 years of education, ≤2 for 0-7 years of education), 3) a Clinical Dementia Rating score of 0.5 or 1.0, and 4) self or partner reported memory complaints. Of the 100 participants initially identified from the ADNI dataset, participants without biomarker data (*n* = 4), with excessive head motion, defined as mean framewise displacement > .15 mm (*n* = 10), and those with poor whole-brain coverage (*n* = 3) were excluded. Data from the remaining 83 participants were used for all analyses.

### CSF Biomarker Assessment and Classification

CSF biomarker concentrations were pulled from the upennbiomk9.rdata file nested in the ADNIMERGE R package. The upennbiomk9.rdata file includes CSF biomarker concentrations from ADNI’s first 3 recruitment waves: ADNI1, ADNI-GO, and ADNI2. P-tau and Aβ_42_ concentrations were measured in picograms per milliliter (pg/mL) by ADNI researchers using the highly automated Roche Elecsys immunoassays on the Cobas e601 automated system. In this study, we used concentrations measured by the Roche Elecsys assay over those measured by the traditional AlzBio3 immunoassay platform as the Roche Elecsys system is the primary assay for ADNI’s current recruitment wave (ADNI3), thus allowing for current analyses and results to be compatible with studies that will utilize future ADNI data releases.

The Elecsys-based assay has a lower and upper measurement limit for Aβ_42_ concentrations (200-1700 pg/ml). Although there were no extrapolated values provided for the lower limit values, extrapolated values for the upper limit, based on calibration curves, and provided by the ADNI group were employed for the current analysis. Of the 83 participants, 13 participants met the upper limit for the Aβ_42_ concentration. For all participants, we computed the ratiometric measure of p-tau/ Aβ_42_, with higher values representing higher pathological state of the two proteinopathies.

### Neuropsychological Measures and Composites

Participants in the ADNI study were administered detailed neuropsychological batteries to examine the following cognitive domains: global cognition, episodic memory, executive functioning, spatial orientation, processing speed, and language. In this study, we focused our analyses on three cognitive domains that are commonly impacted in AD: global cognition, episodic memory, and executive functioning. This *a priori* decision was also driven by the availability of pre-existing, validated composites for the three cognitive domains that sum performance across multiple metrics to yield a summary score.

Measures assessing changes in global cognition tend to be the strongest predictors for conversion to AD (80), and also correlate significantly with clinical symptoms of dementia (81). We employed the Preclinical Alzheimer’s Cognitive Composite Score (PACC) designed to characterize the presence of global cognitive deficits in preclinical AD as one of the cognitive outcome variables in the study. The composite summarizes performance across the domains of episodic memory, executive functioning, and overall cognition, and includes the following measures: Mini-Mental Status Examination total score, Trails-Making Test B, delayed recall score from the Logical Memory II subscale, and the delayed word recall from the Alzheimer’s Disease Assessment Scale– Cognitive Subscale (ADAS-COG). PACC scores were provided in the adnimerge.rdata file for baseline, 6-months, 12-months, and 24-months.

To index changes in episodic memory over a two-year period, we employed the ADNI-MEM composite. This composite includes performance on the Logical Memory I and II tasks, several item scores on the Rey Auditory Verbal Learning Test, the cognitive subscale of the Alzheimer’s Disease Assessment Scale, and the three word recall items from the Mini-Mental State Examination (82). For executive functioning, we employed the ADNI-EF composite developed in the ADNI sample using item response theory. The ADNI-EF composite includes performance on the Digit Symbol Substitution test from the Weschler Adult Intelligence Scale-Revised, Digit Span Backwards Test, Trails-Making A and B, Category Fluency, and Clock Drawing (83). ADNI-EF has been shown to be a stronger predictor of AD conversion than its component measures and has been associated with dichotomized CSF concentrations (83).

ADNI-MEM and ADNI-EF were pulled from the uwnpsychsum.rdata file nested in the ADNIMERGE R package. Although ADNI has longer-term follow-up data, there is significant attrition as a function of the diagnostic group, such that at 36 months follow-up, only 53 MCI individulals had CSF data, neuroimaging, and neuropsychological data and there were no participants with AD with all of these data at 36 months.

### MRI Data Acquisition

Structural and functional MRI data, included in the current study, was acquired using a 3 Tesla Phillips Medical Systems MRI scanner. Each participant’s T1-weighted 3D (TR = 6.7 ms, TE = 3.1 ms, flip angle = 9°, field of view = 256 x 256 x 170-mm^3^, slice thickness = 1.20-mm, voxel size standard = 1.0-mm^3^, voxel size accelerated = 1.11 mm^3^, 170 sagittal slices) was included in the study. A total of 140 volumes of resting-state fMRI data were acquired using the following acquisition parameters: TR = 3000 ms, TE = 30 ms, flip angle = 80°, field of view = 212 x 212-mm^2^, slice thickness = 3.13-mm, voxel size = 3.125-mm^3^, 48 sagittal slices. Five participants had a TR (repetition time) of 2.25 ms. However, we accounted for this discrepancy by using subject-specific TR for all data preprocessing and postprocessing steps that required specifying the TR. All downloaded structural and functional MRI data from the ADNI were first converted from DICOM to NIFTI format and organized according to the Brain Imaging Data Structure (BIDS) (84) and processed through validated quality control pipeline that provide an initial assessment of signal-to-noise (SNR) ratio, framewise displacement (FD), and overall drift rate of signal (85).

### MRI Data Preprocessing

Preprocessing of anatomical and functional data was carried out using fMRIPrep v1.4.1 (86). Each participant’s T1w image was corrected for intensity non-uniformity with N4BiasFieldCorrection (87) and skull-stripped with a *Nipype* implementation of the antsBrainExtraction.sh. Brain surfaces were then reconstructed using recon-all from FreeSurfer v6.0.1(88) and spatially normalized to MNI space through nonlinear registration with antsRegistration tool (89). Brain tissue segmentation of CSF, white-matter (WM) and gray-matter (GM) was performed on the brain-extracted T1w using fast in FSL v5.0.9.

Functional data was slice time corrected using 3dTshift from AFNI (90), motion corrected using mcflirt (91), and co-registered to the corresponding T1w using boundary-based registration (92) with nine degrees of freedom (using bbregister in FreeSurfer v6.0.1). Motion correcting transformations, BOLD-to-T1w transformation, and T1w-to-template (MNI) warp were concatenated and applied in a single step with antsApplyTransforms (ANTs v2.1.0) using Lanczos interpolation. Physiological noise regressors were extracted based on CompCor procedure (93). Several confounding timeseries, including FD and global signal (94), were calculated based on the functional data using the implementation of Nipype. Many internal operations of fMRIPrep use Nilearn (95), principally within the BOLD-processing workflow. For more details of the pipeline see: https://fmriprep.readthedocs.io/en/latest/workflows.html.

Following data preprocessing in fMRIPrep, we excluded the first 3 volumes of each functional data to allow BOLD signal stabilization. We then used *signal*.*clean* in Nilearn (95) to remove the following potential confounding sources of signal variance: mean CSF signal, mean WM signal, mean global signal, and a 24-parameter motion model (i.e. six rigid body motion parameters, six temporal derivatives and their squares,(96)), and linear trend. To further control for potential motion related confounds, we added a spike regressor for each frame with FD value ≥ 0.5 mm (97). Functional data was highpass-filtered in the temporal domain to remove frequency signals less than 0.01*Hz*. No spatial smoothing of functional data was performed (98).

### Whole-Brain Parcellation and Functional Connectivity Estimation

The preprocessed resting-state data for each participant was parcellated into 268 contiguous, functionally defined regions using the Shen parcellation scheme covering the cortex, subcortex, and cerebellum(99). The functional atlas was transformed from MNI space to each participant’s native space. BOLD signal timecourses from each region were extracted by averaging all voxels in that region. The connections between each pair of these brain regions or nodes, defined as edges, were computed by correlating the timecourse of activity between the nodes using Pearson’s correlations. Prior to applying Fisher *z*-transformation to functional connectivity data, we excluded 5 nodes from all subjects as they were missing in 3 or more subjects. These nodes were in the cerebellar (2), prefrontal (1), temporal (1) regions and Brainstem (1), resulting in a 263×263 FC matrix for each participant, with cells of the matrix representing the magnitude of these edges or correlations.

### Connectome-Based Predictive Modeling

#### Derivation of PATH-fc functional connectivity model

Figure 1 presents the schematic representation of the various steps of CPM. First, this model employs a leave-one-out-cross-validation (LOOCV) procedure, wherein the data is divided into a training set (n-1 participants) and a test set, with this process repeated until each participant has served as the test participant (Figure 1A). To define an AD pathology network, for all participants in the training set, each cell of the functional connectivity matrix was correlated with p-tau/Aβ_42_and thresholded for significant correlations, across participants, at *p*<.01 (Figure 1B), resulting in a set of edges where each surviving edge is either positively or negatively associated with CSF-biomarker accumulation (Figure 1C). To further reduce the effect of motion, we regressed out the mean FD values for each participant at this edge-selection step. As noted above, we only examined the high pathology network showing edges positively correlated with CSF-biomarker accumulation. This CPM technique thus yields predicted AD pathology scores for each participant (Figure 1D), which are then correlated using Spearman’s rank correlation (*ρ)* with observed scores to generate a measure of model performance (Figure 1E). Permutation testing, involving shuffling observed AD pathology scores and implementing CPM LOOCV 1000 times to build a null distribution, was employed to evaluate if the prediction performance for the PATH-fc model was significantly better than expected by chance. We also ran permutation testing for each validation analysis described below.

#### Application of PATH-fc model to predict cognitive decline

Changes in cognitive outcomes (PACC, ADNI-MEM, and ADNI-EF) over time were modeled using linear mixed effects models. We estimated the rate of change over two years by modeling performance at baseline, 6-months, 12-months, and 24-months using a mixed model that included fixed effects of time (linear), diagnosis group, and their interaction. From this model the empirical best linear unbiased predictor (eBLUP) for each subject was estimated and used as the subject-specific measure of cognitive decline. In order to determine whether our AD pathology network could also be used to predict rate of cognitive decline in an unseen individual, we employed the averaged connectivity strength within the PATH-fc high network to train three predictive models for rate of cognitive decline on the PACC, ADNI-MEM, and ADNI-EF, each evaluated using a LOOCV approach.

#### Anatomical and Network Localization of PATH-fc Model

Edges were deemed reliable if they appeared in every round of LOOCV iterations across all subjects (39). The final mask, comprising of edges present in every round of the LOOCV, represented the PATH-fc model with 581 edges distributed across all macroscale regions. To summarize the involvement of the macroscale regions as well as the canonical networks, we parcellated the 268 nodes of the Shen atlas based on their spatial overlap with the 10 macroscale regions identified in Finn et al., (100) and the 10 canonical networks identified in (101) (Figure S1). For the macroscale regions, the following anatomical labels were employed: prefrontal, motor, insular, parietal, temporal, occipital, limbic, cerebellum, brainstem, and subcortical. The canonical networks included the following: medial frontal, frontoparietal, default mode, motor, visual I, visual II, visual association, salience, subcortical, and cerebellar networks. For the canonical networks, we summarize the overall involvement of the various networks, but also the relative involvement of each network, adjusted for size (or number of nodes) of the network in both the Shen atlas and the PATH-fc model (39). And, finally, we calculated degree – sum of the number of edges – for each node of the PATH-fc model to highlight nodes that show the highest number of connections with other nodes in this AD pathology-based model. Ring plots were created using Flourish data visualization tools (https://flourish.studio/). BrainNet Viewer toolbox (102) was used for brain-based visualization of the PATH-fc.

#### Validation Analyses: Choice of feature selection threshold

As the choice of edge threshold selection is inherently arbitrary, we tested the sensitivity and robustness of the PATH-fc model to additional thresholds with *p*-values of 0.1, 0.05, 0.005, and 0.001 (40). *Alternative Parcellation Scheme*: Previous studies have shown that the specific choice of a parcellation scheme may impact the results from brain network analysis (103, 104). To investigate potential parcellation effects, we employed an additional functional atlas consisting of 360 brain regions (Glasser360, (105)). Similar to the main analysis, we estimated mean BOLD timecourses and excluded four nodes (two each in the prefrontal and temporal regions) from all subjects as they were missing in three or more subjects. This resulted in a 356×356 FC matrix for each subject. All analyses described under connectome-based predictive modeling were repeated, including derivation of the PATH-fc model, and application of the PATH-fc model to predict AD pathology and change in cognitive decline.

## Data Availability

The current study is a secondary analysis of publicly available data from The Alzheimer Disease Neuroimaging Initiative (ADNI). All data is publicly available.

## Acknowledgments

The project described in this publication was supported by Award Number UL1TR002733 from the National Center For Advancing Translational Sciences and Award Number R01AG054427 from the National Institute On Aging of the National Institutes of Health. The content is solely the responsibility of the authors and does not necessarily represent the official views of the National Center For Advancing Translational Sciences, the National Institute on Aging or the National Institutes of Health.

## Supplementary Tables and Figures

**Table S1.**
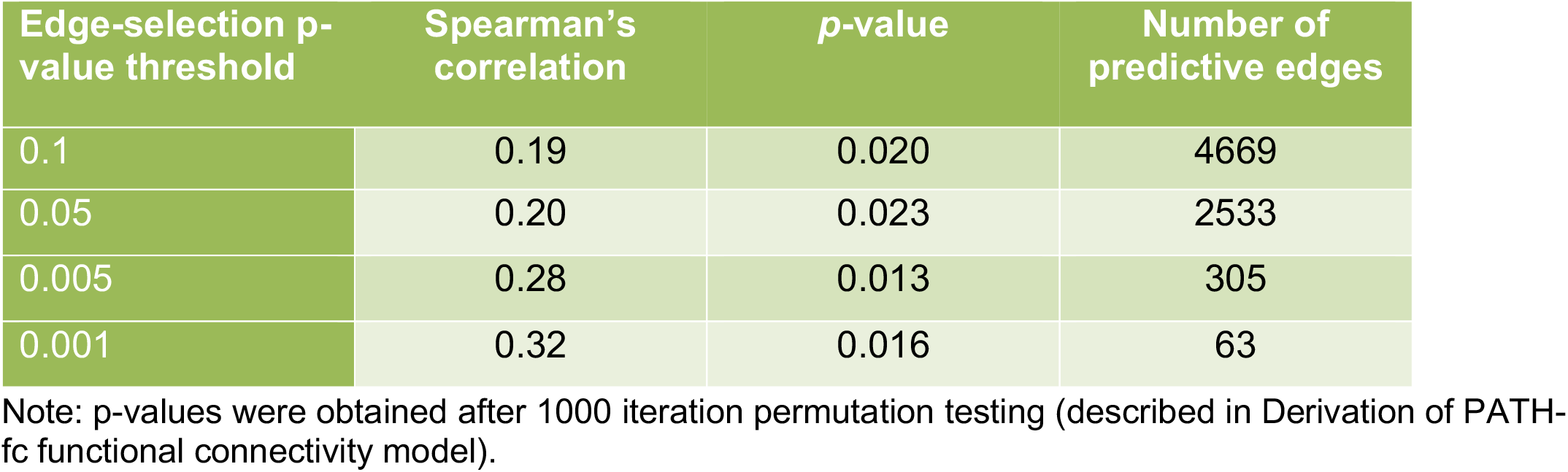
Internal Validation Prediction Results at Various Edge Selection Thresholds. High pathology model performance was comparable across various edge-selection thresholds.

**Figure S1:**
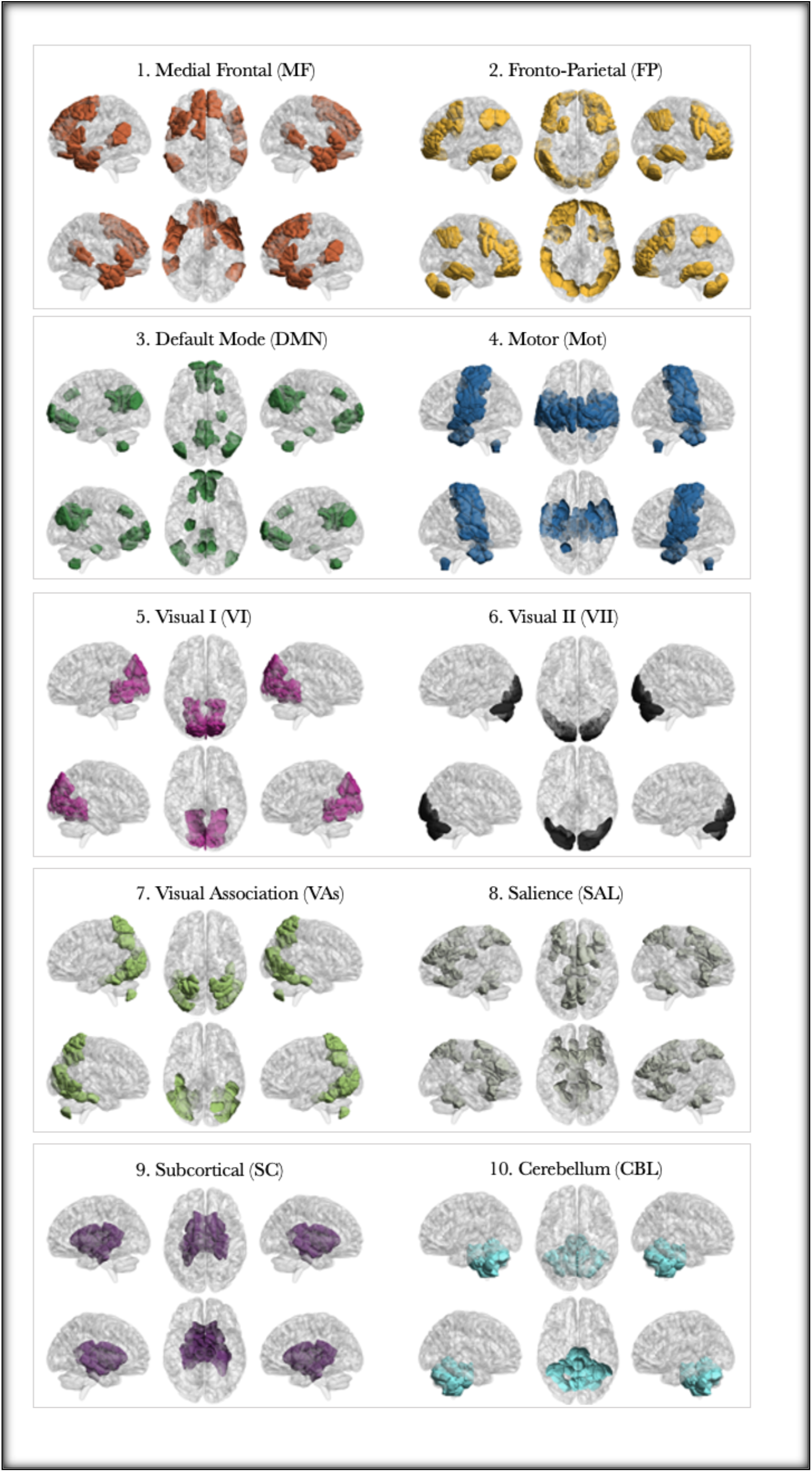
Presents the 10 canonical networks used for network localization of the PATH-fc model.

**Figure S2:**
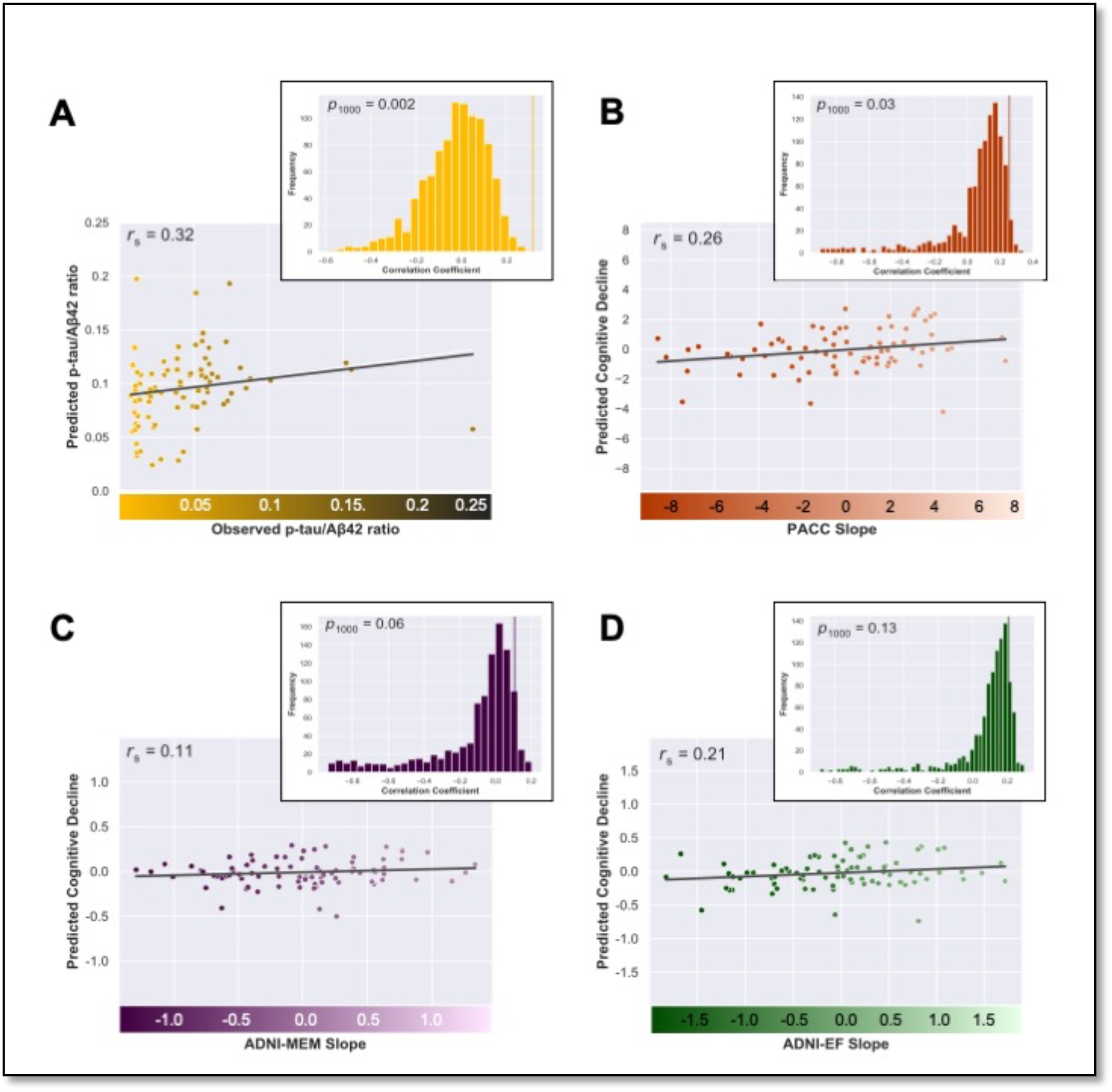
Results of the internal model fit of the PATH-fc model generated using the Glasser atlas. Internal model fit is shown for p-tau/AB42 ratio (A), and declines in PACC scores (B), ADNI-MEM (C), and ADNI-EF (D). The scatterplots represent Spearman correlation between observed scores (AD pathology, PACC, ADNI-MEM, and ADNI-EF) and predicted scores for the same metric derived using cross-validation. Histogram represents the randomly shuffled connectivity matrices and respective metric pairings over 1000 iterations to compute the final p-value for the significance.

**Figure S3:**
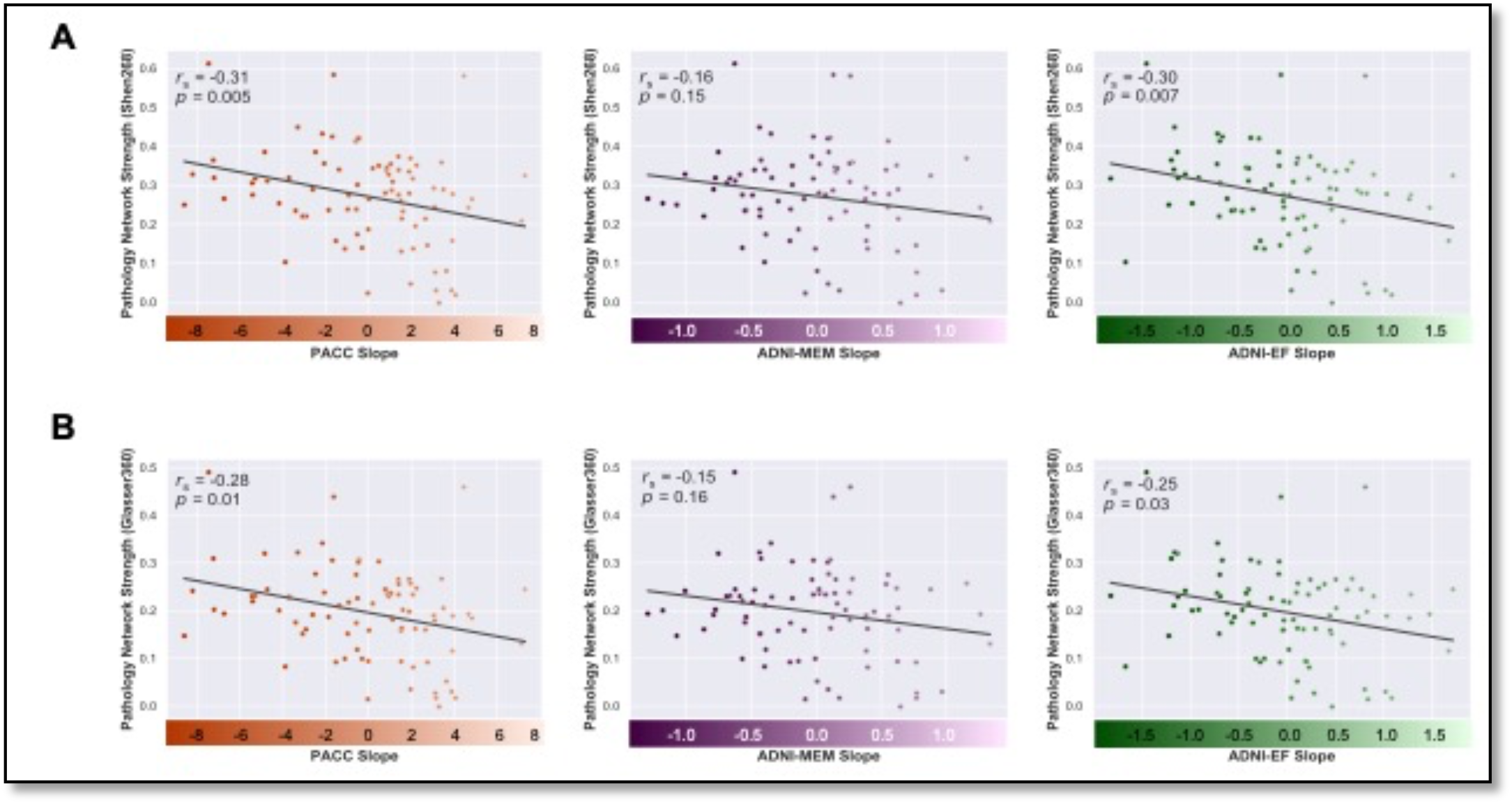
Panel A shows the correlation between network strength of the PATH-fc model and decline in composite scores for the three domains of cognition. The PATH-fc model was derived using the parcellation scheme of the Shen atlas. To ensure that our model was robust to the choice of atlas, we also replicated the analyses using the Glasser atlas. Panel B presents the results from the Glasser atlas derivation, with the scatter plots showing a correlation between network strength of the PATH-fc model and decline in cognitive functioning.

